# Computing the Prevalence and Severity of an Epidemic Using only the Distribution of Simple Tests for Infection Confirmation

**DOI:** 10.1101/2021.05.04.21256588

**Authors:** Yuval Shahar, Osnat Mokryn

## Abstract

Epidemics and Pandemics such as COVID-19 require estimating total infection prevalence. Accurate estimates support better monitoring, evaluation of proximity to herd immunity, estimation of infection fatality rates (IFRs), and assessment of risks due to infection by asymptomatic individuals, especially in developing countries, which lack population-wide serological testing.

We suggest a method for estimating the infection prevalence by finding the *Pivot group*, the population sub-group with the highest susceptibility for being confirmed as positively infected. We differentiate *susceptibility to infection*, assumed to be uniform across all population sub-groups (a key assumption), from *susceptibility to developing symptoms and complications*, which differs between sub-groups (e.g., by age). We compute the minimal infection-prevalence factor by which the number of positively confirmed patients should be multiplied that allows for a sufficient number of Pivot-group infections that explains the number of Pivot group confirmations.

We applied the method to the COVID-19 pandemic, using UK and Spain serological surveys. Our key assumption held, and actual infection-prevalence factors were consistent with our predictions. We computed minimal infection-prevalence factors, and when possible, assessed IFRs and serology-based IFRs, for the COVID-19 pandemic in eight countries.

Estimating a lower bound for an epidemic’s infection prevalence using our methodology is feasible, and the assumptions underlying it are valid. The use of our methodology is often necessary for developing countries, especially in the early phases of an epidemic when serological data are not yet available or when new mutations of a known virus appear.

## Introduction

A common problem when attempting to manage epidemics and pandemics is assessing the total infection prevalence of the disease, which was often a key issue with the COVID-19 pandemic. The problem is often referred to as assessing the total “*Infected Iceberg’s*” size (including the portion of the Iceberg that is “underwater,” which is composed of asymptomatic infected individuals)^1–5^. Correct estimation of the total infection prevalence also bears directly on the *infection fatality rate* (IFR); a good *lower bound* for the first estimate provides, indirectly, a good *upper bound* for the second estimate.

One suggestion to solve the problem is to use serological testing of the population, preferably measured randomly, to assess the overall infection prevalence or Iceberg size^6–10^. For example, in the case of the COVID-19 pandemic within Spain, using the serology has led to a mean of 5% positive seroprevalence using point of care (PoC) testing and a mean of 4.7% positive seroprevalence using a laboratory-based immunoassay testing^11^. In the case of COVID-19 in the UK, a large-scale self-administered immunoassay with over 100,000 volunteers had suggested that by the time in which the serological tests were performed, a mean of 6.4% of the population had been infected^12,13^.

However, in general, serological and antibody home testing often have a known caveat, since previously symptomatic people might be more likely to participate in these tests^14^. Another caveat is that they may be less reliable with time in the case of a decline of neutralizing antibody responses with time^15^. Furthermore, serological tests are often difficult and costly to administer, especially in developing countries^16–18^.

An alternative strategy for determining infection prevalence is the performance of massive acute disease testing during an epidemic. In the case of the COVID-19 pandemic one such strategy that was suggested, which attempts to reduce costs, is *pooled testing*^19^. However, pooled testing requires a dedicated testing infrastructure and the overcoming of multiple technical hurdles. Other researchers have assessed through simulation the effect of various assumptions on the proportion of asymptomatic cases and their infectivity and compared the results to actual data^20^.

Here, we suggest a simple statistical method that uses only the distribution of the data of the patients who are confirmed as positive for the disease in question, for setting a lower bound on the size of an epidemic’s Iceberg (and correspondingly, an upper bound on the IFR).

## Method

Our method for estimating the minimal Iceberg size relies on finding a sub-group in the population for which the relative risk for being positively confirmed as infected is the highest. We refer to this high-risk sub-group as the *Pivot group*. We define the *Iceberg Factor* (IF) as the ratio of the total size of the infected population, to the number of confirmed infected individuals. We further define the *Minimal Iceberg Factor* (MIF) as the smallest IF that explains the number of individuals in the Pivot group.

A key assumption in our method is the *infection-uniformity assumption*: The risk of initial infection, which we refer to as *S*_0_, is uniform across all population sub-groups; to compute a valid lower bound on the IF, it is enough that *S*_0_ should at least not be greater for the Pivot sub-group. That is, we assume that the initial infection process is a random stochastic process, and thus, the proportion of each infected sub-group within the total infected population is similar to its proportion in the overall population. We differentiate the probability of initial infection, *S*_0_, from the conditional probability of being symptomatic given that the patient is infected, *S*_1_, which, in the specific case of COVID-19, is known to be age-related. Thus, even though the initial infection probability *S*_0_ is similar across all sub-groups, such as different age groups, some sub-groups might well be over- or under-represented within the group of patients confirmed as *positive*. For example, the elderly sub-group might be over-represented in the positively confirmed group of COVID-19 patients in spite of a uniform S_0_, because elderly patients have a higher *S*_1_, are more likely to be symptomatic after being infected, and thus more likely to be confirmed as positive. (The Results support our assumptions).

In this study, we focus on applying the method to the COVID-19 pandemic and only for demographical sub-groups, specifically, age-related subgroups. In general, this focus can be broadened, and other sub-groups, such as defined by gender or ethnicity, might be used in the analysis. As we shall see when presenting our algorithm, the MIF is in fact the relative risk (*Lift*) of the Pivot sub-group. Thus, **given the infection-uniformity assumption (a uniform** *S*_0_**), this IF is the minimal one that can explain the existence of all of the Pivot sub-group members that were confirmed as positive**.

However, the MIF, and the respective overall Infected Iceberg size, might be *smaller*, if, by chance, *more* people from the Pivot sub-group within the overall population were “sampled” by the random infection process. Thus, we also need to test whether a sufficiently large number of people from the Pivot sub-group (specifically, the number that is needed to explain their existence within the known positively-confirmed group) might have been sampled from the overall population in a reasonably likely manner (i.e., in a statistically insignificant fashion), even when given a smaller Iceberg Factor, and thus a smaller overall Iceberg size.

Therefore, we test the reasonable likelihood of each potential Infected Iceberg size (corresponding to a given IF) by applying a proportion test to find whether the proportion of the Pivot sub-group in the Infected Iceberg of a particular country might be, purely by chance, sufficiently higher than their proportion within the country’s overall population, so as to explain the actual confirmed positive numbers of that Pivot sub-group, but still be larger only in a statistically insignificant fashion. Thus, in our study, in addition to computing the MIF, we computed the smallest MIF that still explain the number of Pivot Group members that are confirmed as positive, but for which the assumption of a “reasonably likely” sampling process due to the infection is not rejected, which we refer to as the *Statistically Insignificant Minimal Iceberg Factor* (**SIMIF**). That is, the SIMIF is the minimal IF for which the proportion test (for the Pivot sub-group’s proportion within the Infected Iceberg) was still insignificant.

Our suggested method is as follows:

1. Split the population into disjoint exhaustive sub-groups. For example, by age, gender, or both.
2. Find the *Pivot sub-group*, the population sub-group that displays the maximal relative-risk (*Lift*) for being positively confirmed as infected. This is the sub-group for which its proportion within the confirmed (positive) infected patients, compared with its percentage in the population, is the highest.
3. Given the Key Assumption, and thus assuming that the distribution of groups within the infected population is similar to their population distribution, set the MIF to be the Lift of the Pivot sub-group. Thus, the resulting infected Iceberg includes enough members of the Pivot group. Note that the infection-uniformity assumption can be relaxed to the assumption that the infection rate of the Pivot sub-group is not greater than that of the rest of the population, to maintain the MIF as a lower bound on the IF.
4. To allow for statistical deviations, compute the MIF that, even allowing for an insignificant statistical deviation from the Pivot group’s proportion in the population during the infection process, might still contain a sufficient number of the Pivot group members to explain the number found in the “visible” part of the Iceberg. That is the *Statistically Insignificant Minimal Iceberg Factor* (SIMIF).

Given the MIF, which is a *lower bound* on overall infection prevalence, we compute the *upper bound* on the *Infection Fatality Rate* (IFR), by dividing the number of deaths due to the disease by the size of the estimated Infected Iceberg (i.e., the number of positively confirmed cases multiplied by the MIF).

In the Results Section, we demonstrate in detail the application of this method to the COVID-19 pandemic, using data from two countries (the UK and Spain) and then summarize the results for a total of eight different countries; in one of them (UK) we performed the computation for two data sets acquired at different time points (June and September 2020), and in another case (USA) we performed the computation for data sets acquired at two different time points from two different regions (New York City and New York State).

Table 1 summarizes each country’s aggregated information of the PCR-RT COVID-19 Confirmed individuals and at which date, the source from which the data was obtained, and the country’s population. We analyzed only secondary data available in the public domain, with no need for approval by the ethics committee in research.

**Table 1.** Covid-19 PCR-RT positive test results data used in this research

| **Country** | **Date** | **Source** | **PCR-RT COVID-19 Confirmed individuals** | **Country population<sup>34</sup>** |
| --- | --- | --- | --- | --- |
| **Spain** | May 21 2020 | Spanish Centro de Coordinación de Alertas y Emergencias Sanitarias<sup>20</sup> | 252,283 | 46,736,782 |
| **UK** | June 10 2020 | UK Department of Health and Social Care Statistical Bulletin<sup>22</sup> | 222,441 | 67,530,161 |
| **UK** | Sep. 1 2020 | UK Department of Health and Social Care Statistical Bulletin<sup>25</sup> | 287,389 | 67,530,161 |
| **USA NY State** | March 30 2020 | US dept. of Health<sup>26</sup> | 117,522 | 19,542,209 |
| **USA NYC** | May 16 2020 | NYC dept. of Health<sup>27</sup> | 357,230 | 8,336,817 |
| **Italy** | June 24 2020 | *Prodotto dall'Istituto Superiore di Sanità (ISS)*<sup>28</sup> | 238,042 | 60,359,546 |
| **Norway** | June 19 2020 | The Norwegian Institute of Public Health<sup>29</sup> | 8,708 | 5,378,859 |
| **Sweden** | Dec. 16 2020 | Statista, Sweden<sup>30</sup> | 357419 | 10036391 |
| **Belgium** | July 2 2020 | Belgian Institute for Health<sup>31</sup> | 61,507 | 11,539,326 |
| **Israel** | July 4 2020 | Israel Government Data Repository<sup>32</sup> | 28,259 | 8,519,372 |

## Results

We shall first demonstrate the value and outcomes of our methodology using the COVID-19 RT-PCR data for Spain on May 22, 2020^21^. At that point,*C*_*pos*_= 252,283 positive (confirmed) cases were known, as can be seen in Table 2. The table further depicts the number and percent of individuals in each age group out of the country’s population, and the number of confirmed (PCR-RT COVID-19 Confirmed individuals) and percent out of all confirmed of individuals in each age-group.

**Table 2.** Number and age distribution of Spanish and UK citizens within the overall population and within the confirmed COVID-19 cases, for Spain on May 22, 2020, and for the UK on June 10, 2020.

|  | SPAIN |  |  |  | UK |  |  |  |
| --- | --- | --- | --- | --- | --- | --- | --- | --- |
| Age | # in pop. | % from pop. | # of confirm | % from confirm | # in pop. | % from pop. | # of confirm | % from confirm |
| 0-9 | 4,340,417 | 9.29% | 998 | 0.4% | 8,065,283 | 11.95% | 1,900 | 0.85% |
| 10-19 | 4,682,339 | 10.02% | 1,861 | 0.74% | 7,569,160 | 11.21% | 4,060 | 1.83% |
| 20-29 | 4,652,133 | 9.95% | 14,562 | 5.77% | 8,630,614 | 12.78% | 23,728 | 10.67% |
| 30-39 | 6,158,281 | 13.18% | 24,075 | 9.54% | 9,203,569 | 13.63% | 27,895 | 12.54% |
| 40-49 | 7,935,505 | 16.98% | 36,872 | 14.62% | 8,624,679 | 12.77% | 30,643 | 13.78% |
| 50-59 | 6,944,643 | 14.86% | 44,591 | 17.67% | 9,138,365 | 13.53% | 36,343 | 16.34% |
| 60-69 | 5,200,462 | 11.13% | 35,713 | 14.16% | 7,206,475 | 10.67% | 23,196 | 10.43% |
| 70-79 | 3,921,750 | 8.39% | 33,814 | 13.4% | 5,673,457 | 8.40% | 24,304 | 10.92% |
| 80+ | 2,901,252 | 6.21% | 59,797 | 23.7% | 3,418,559 | 5.06% | 50,372 | 22.64% |
| All | 46,736,782 | 100% | 252,283 | 100% | 67,530,161 | 100% | 222,441 | 100% |

Consider the Spanish age distribution of the confirmed cases. Out of *C*_*pos*_ = 252,283 positive cases, the number of 80 years or older cases, *C*_*pos*.80+_, was 59,797 (23.7%) – 3.82 as much as their proportion in the Spanish population^22^, *POP*_*prop*.80+_ which is only 6.21% (2,901,252 of 46,736,782). This sub-group has the highest relative risk (*Lift*) for being confirmed as positive. Thus, the sub-group of 80+ year old people is the Spanish Pivot sub-group, and its Lift is 3.82. Thus, 3.82 would be the MIF for Spain at that point in time.

In other words, at least 963,721 people must have already been infected at that point in time in Spain, to explain the number of positively confirmed cases from its Pivot sub-group.

When we follow the same procedure for the United Kingdom using its June 10, 2020 data^23^ (see Table 2), the minimal Iceberg size that explains the number of positive confirmed cases in the UK’s Pivot, or highest-risk, sub-group, the 80+ years old age-group (4.68% of the British population^24^) at that point in time, *C*_*pos*.80+_ = 50,372, must be at least their relative risk for being confirmed as positive, namely, 4.48 the number of total positive cases found at that time (*C*_*pos*_= 222,441). Thus, The UK MIF on June 10th, 2020 was 4.48.

Therefore, a total of at least *C*_*tot*_ = 1,112,205 British people must have been infected at that point in time, most of them being “underwater” (unconfirmed), to explain the finding at that time of 50,372 positive cases in the 80 + years old age group.

However, based on statistical reasoning, another option might be suggested to explain the number of positively confirmed cases from the Pivot sub-group in Spain or in the UK, using a smaller IF, but without leading to a smaller number of positively confirmed patients from the Pivot sub-group. Perhaps the proportion of infected 80 + years older adults in the Iceberg was, by chance, higher than their proportion within the population (even assuming that the likelihood of infection does not depend on age); and somehow, all of the infected older adults were tested and found positive. Could that explain the number of positively confirmed octogenarians while using a smaller IF, namely, a smaller Infected Iceberg?

In the case of the Spanish example, note that if the Iceberg’s age distribution is similar to that of the Spanish population, it would contain, for an Iceberg Factor of 3.0, only *I*_80+_= 46,982 cases, and thus we are short of 12,815 positive patients in that age group. But perhaps the proportion of infected 80 + years older people in the Spanish Iceberg was, by chance, higher than their proportion within the Spanish population?

To explore this explanation, we applied a proportion test to see whether it is reasonable that, given the proportion of the 80 + years old population in Spain, enough positive cases might have existed at random within the Spanish Iceberg. That is, whether the 2,935,720 people who are 80 + years old, out of Spain’s population of 62,676,180 citizens (i.e., 6.21%), might have randomly produced, through the “random sampling” of being infected, the minimal necessary number of 59,797 positive cases, within an only threefold (i.e., IF = 3.0) Iceberg size of 756,849 (i.e., 7.9%), assuming an age-oblivious infection process.

The result is: z-statistic = 60.92103; Significance level *p* < 0.0001; 95% CI of observed proportion: 7.84% to 7.96%. (Compare this confidence interval to Spain’s 80 + years age group, which includes only 6.21% of the population). Thus, the Null Hypothesis is rejected at enormous odds. Thus, the IF is highly likely to be larger than three times the total number of confirmed positive cases to explain the number of confirmed cases in the 80+ years age group. In fact, any IF ≤ 3.76 would result in rejecting the null hypothesis at a level of significance greater than *p* < 0.05. So the Spanish SIMIF at that point was 3.77.

For the British data and an example factor of four, the results are similar: z-statistic 43.76; Significance level *p* < 0.0001; 95% CI of observed proportion: 5.61% to 5.71%. (Compare this confidence interval to the UK’s 80 + years age group, which includes only 4.68% of the population). Thus, the British IF then must have been larger than four. In fact, for the UK on June 10th 2020, any IF ≤ 4.43 would result in rejecting the null hypothesis at a level of significance greater than *p* < 0.05, so the UK SIMIF at that point was 4.44. Since not all infected cases were confirmed as positive, both the Spanish and the UK Icebergs must have been larger.

We followed this procedure for multiple countries or large regions whose data, mostly during the early COVID-19 pandemic phase, were available (Spain^25^, the UK at two different time points^23,26^, New York State^27^ (USA), New York City^28^ (USA), Italy^29^, Norway^30^, Sweden^31^, Belgium^32^, Israel^33^). For each of them, we established the minimal lower bound on the Iceberg factor that explains the population age-based distribution, assuming an age-independent S0. The lower bound ranged from 1.35 (NYC) to 5.1 (Belgium). The results are summarized in Table 3, showing for each country the date the data was collected; the number of individuals that were PCR-RT positive, termed Tested Positive; the age-group of the Pivot group for which the relative risk is higher, their percentage in the population and their relative risk. Then we show the MIF calculated from the Pivot group information, and the Iceberg size that corresponds to the MIF, for each country. We continue to show the result of the numerical calculations by depicting the SIMIF and its corresponding Iceberg size, and the results of the Proportion test for the SIMIF for the Pivot group (z-stat, p-value, and confidence interval). The last column shows for each country and data date the Interval within which the SIMIF becomes significant.

**Table 3.** Minimal Iceberg Factor (MIF), Statistically Insignificant Minimal Iceberg Factor (SIMIF), and proportion test calculations for the COVID-19 pandemic Pivot groups in eight different countries, for an overall ten different dates

| Country |  | Spain | USA NYS | USA NYC | UK | UK | Italy | Norway | Sweden | Belgium | Israel |
| --- | --- | --- | --- | --- | --- | --- | --- | --- | --- | --- | --- |
| MM.DD.YY |  | 5.22.20 | 3.31.20 | 5.16.20 | 6.10.20 | 9.1.20 | 6.24.20 | 6.19.20 | 12.16.20 | 7.2.20 | 7.4.20 |
| Tested Positive |  | 252,283 | 117,522 | 357,230 | 222,441 | 287,389 | 238,042 | 8,708 | 357,419 | 61,507 | 28,259 |
| Pivot group(Covid-19 age group) |  | 80+ | 50-64 | 55-64 | 80+ | 80+ | 80+ | 50-59 | 40-49 | 80+ | 20-29 |
| Pivot % of pop. |  | 6.21% | 20.01% | 11.98% | 5.06% | 5.06% | 7.17% | 13.04% | 12.73% | 5.71% | 13.84% |
| relative Risk of Pivot for pos. |  | 381.83% | 141.31% | 134.54% | 447.33% | 379.74% | 353.21% | 142.39% | 142.44% | 509.98% | 158.81% |
| MIF | MIF | 3.82 | 1.42 | 1.35 | 4.47 | 3.8 | 3.55 | 1.43 | 1.43 | 5.1 | 1.6 |
|  | Total Implied Iceberg size | 963,721 | 166,881 | 482,261 | 996,536 | 1,092,078 | 845,049 | 12,452 | 511,109 | 313,686 | 45,214 |
| SIMIF Proportion Test | SIMIF | 3.76 | 1.39 | 1.33 | 4.43 | 3.76 | 3.5 | 1.34 | 1.41 | 5.02 | 1.6 |
|  | Total Implied Iceberg size | 953,630 | 165,356 | 475,115 | 985,414 | 1,080,583 | 833,147 | 11,669 | 503,961 | 308,765 | 43,801 |
|  | Pivot proportion from SIMIF Iceberg | 6.27% | 20.34% | 12.12% | 5.11% | 5.11% | 7.24% | 13.86% | 12.86% | 5.80% | 14.18% |
|  | z-stat | 2.428 | 3.334 | 2.972 | 2.265 | 2.371 | 2.477 | 2.63 | 2.769 | 2.155 | 2.061 |
|  | p-value | 0.0152 | 0.0009 | 0.003 | 0.0235 | 0.0177 | 0.0133 | 0.0085 | 0.0056 | 0.0311 | 0.0393 |
|  | 95% CI | 6.22%-6.32% | 20.15%-20.541% | 12.03%-12.21% | 5.07%-5.15% | 5.07%-5.15% | 7.18%-7.3% | 13.24%-14.5% | 12.77%-12.95% | 5.72%-5.88% | 13.83%-13.84% |
| Interval within which SIMIF becomes significant |  | [3.76,3.77] | [1.39,1.40] | [1.33,1.34] | [4.43,4.44] | [3.76,3.77] | [3.5,3.51] | [1.34-1.35] | [1.41-1.42] | [5.02-5.03] | [1.55-1.56] |

All that remains now is to validate our assumption that the initial infection susceptibility S_0_ is indeed age-invariant, and in particular, not significantly higher for our Pivot group, which in this case consists of the older people. We can easily validate this assumption by examining serological testing results from Spain and the UK (Table 4), depicted by Age group and the type of test. In the Spanish population, Blood samples were taken during April 27 to May 11, from 61,075 participants who received a point-of-care antibody test; if they agreed, a more definitive chemiluminescent microparticle immunoassay was also performed. The mean portion of older adults demonstrating evidence for previous COVID-19 infection was quite similar, considering both test types, to the portion of seropositive cases within the other age groups. In the case of the laboratory-based immunoassay, it is even lower than that portion within all other age groups, except for children and adolescents. Serological tests in the UK were performed during June 20 to July 136, using a self-administered lateral flow immunoassay (LFIA) test for IgG among a random population sample of 100,000 adults over 18 years. The results certainly do not suggest a higher infection-susceptibility risk, *S*_0_, for the elderly population: The portion of 75 + years old adults demonstrating evidence in their blood samples for previous COVID-19 infection was the lowest of all age groups for which the test was performed, thus further validating our assumption. In both countries, the actual Iceberg factors computed from the serological tests (9.32 by PoC or 8.49 by immunoassay for Spain, and 17.00 for the UK) were, as predicted, considerably higher than the MIF lower bound computed by our method (3.82 and 4.48) for chronologically similar periods, and certainly higher than the SIMIF. The serology tests in New York State from March 2020^34^ yields an Iceberg Factor of 22.23.

**Table 4.** Serology results for COVID-19 in Spain from April 27 to May 11, using two different serological tests, and in the United Kingdom from June 20 to July 13, using lateral flow immunoassay (LFIA). Estimates of prevalence adjusted for imperfect test sensitivity and specificity; 95% Confidence Interval is specified for each estimate

| SPAIN<br>Relative frequency of positives (%) |  |  | UK<br>Relative frequency of positives (%) |  |
| --- | --- | --- | --- | --- |
| Age | Point-of-care test | Immunoassay | Age | LFIA |
| 0-19 | 3.4 [2.9-3.9] | 3.8 [3.2-4.6] | 18-24 | 7.1 [6.5-7.8] |
| 20-34 | 4.4 [3.7-5.1] | 5.0 [4.3-5.8] | 25-34 | 7.0 [6.5-7.4] |
| 35-49 | 5.3 [4.7-5.9] | 4.9 [4.3-5.5] | 35-44 | 5.7 [5.3-6.0] |
| 50-64 | 5.8 [5.3-6.5] | 4.7 [4.1-5.3] | 45-54 | 6.1 [5.8-6.4] |
| >64 | 6.0 [5.4-6.8] | 4.5 [3.8-5.3] | 55-64 | 5.5 [5.2-5.9] |
|  |  |  | 65-74 | 3.7 [3.4-4.0] |
|  |  |  | 75+ | 3.6 [3.2-4.1] |
| Spain Factor by Serology: 4.58 - 5.033 |  |  | UK Factor by Serology: 5.25 - 5.96 |  |

The MIF and the SIMIF were quite close in the cases we analyzed in detail. We proceed to compute the upper bound on the IFR. Recall that the MIF is a lower bound on the Iceberg Factor. Then, the estimated Infected Iceberg size is the number of positively confirmed cases multiplied by the MIF. We can compute an upper bound on the IFR by dividing the size of the estimated Infected Iceberg by the fatalities from the infection. Given the COVID-19 disease data, we chose the fatality rate date to be two weeks later than the date of the Serology test.

The IFR upper bound computed by the lower bound provided by the MIF in Spain (given the number of deaths by June 6th, 2020, two weeks after May 22, 2020) was 3.03%, while the IFR computed by serology test results (which can be considered as closer to the true IFR) was 1.24% (PoC) or 1.36% (immunoassay); for the UK, the IFR upper bound computed by the lower bound provided by the MIF was 4.04%, while the IFR computed by serology was 1.06%; for New York State, the IFR upper bound computed by the lower bound supplied by the respective MIF was 9.29%, while IFR computed from the serological test results was 0.59%. The full details appear in Table 5, showing for countries for which we have serological data, the date at which the serology data was reported and the serology test type, the COVID-19 fatality rate date, and the corresponding death toll at that date, The Serology-based Iceberg Factor and Size, and the calculated IFR according to the Serology and according to the MIF.

**Table 5.** Upper bounds on the infection fatality rate (IFR) of covid-19 for countries in which serological test results were available, calculated through the minimal iceberg factor (MIF) and through the actual serological test results

| Countr<br>y | Serology<br>type | Serology<br>date | Covid<br>-19<br>death<br>toll<sup>35</sup> | Covid-19<br>death toll<br>date | Serology<br>-based<br>Iceberg<br>Factor | Serology<br>-based<br>Iceberg<br>Size | Serology<br>-based<br>IFR % | MIF_base<br>d Upper<br>bound on<br>IFR % |
| --- | --- | --- | --- | --- | --- | --- | --- | --- |
| Spain | PoC | 5.11.202<br>0 | 2920<br>4 | 6.6.2020 | 9.32 | 2,352,26<br>2 | 1.24 | 3.03 |
| Spain | Immunoassa<br>y | 5.11.202<br>0 | 2920<br>4 | 6.6.2020 | 8.49 | 2,141,99<br>3 | 1.36 | 3.03 |
| UK<br>June | LFIA | 7.13.202<br>0 | 4116<br>7 | 7.27.202<br>0 | 17.00 | 3,781689 | 1.06 | 4.04 |
| NYS<br>US | Immunoassa<br>y | 3.30.202<br>0 | 1550<br>0 | 4.15.202<br>0 | 22.23 | 2,612,26<br>8 | 0.59 | 9.29 |

## Discussion

Estimating at least a lower bound for the total number of the infected cases in a given population is key to managing an epidemic, and certainly a pandemic. Among other benefits, it supports an assessment of the risk due to asymptomatic cases, and the creation of a more realistic upper bound on the IFR. Forming an estimate without serological testing is especially important when it is costly and difficult to administer them, as is often the case in developing countries.

The line of reasoning suggested here, based on finding the highest-lift Pivot group (we happened to use the age sub-groups), provides a solid lower bound for the size of an Infection Iceberg at any point in time. We have ignored, in the case of the COVID-19 pandemic, the RT-PCR sensitivity and specificity, but we assume they do not vary across age groups; the number of confirmed cases is by itself only a lower bound, due to the PCR’s limited sensitivity. Our methodology can be used in the early phases of any pandemic when serological data are not yet available, and when vaccines are not available; or when new mutations of a known virus appear, which are resistant to an existent vaccine; or when a new virus [to which a vaccine does not exist] is detected, to monitor a pandemic and to compute the IFR’s upper bound.

The MIF is only a lower bound: Only a portion of the Pivot sub-group’s infected members are likely to be confirmed as positive. Furthermore, the infection-uniformity assumption can be relaxed: As long as the Pivot sub-group was not infected at a relatively *higher* rate, the estimated MIF is a valid lower bound for the serology-based IF.

There are some limitations to our methodology. In particular, it is useful when positively confirmed cases are detected mostly due to a symptomatic presentation by the patients (which is governed by the symptomatic-susceptibility probability S_1_), as was common during the early phase of the COVID-19 pandemic, or when some underlying process creates a high variability between different sub-groups, regarding the probability of being positively confirmed. It is less useful when positively confirmed cases are detected at random, such when a general screening of the population is performed (whose results are governed by the infection-susceptibility probability S_0_). The later situation became more common during the more advanced phases of the COVID-19 pandemic, as the number of tests grew, and the indications for performing them had expanded.

The MIF might add insights to pandemic-related differences across different countries and times. Thus, in the case of the COVID-19 pandemic, in September, the Lift for the UK 80+ age group *decreased* compared to its value in June, possibly reflecting a greater cautiousness of the older UK population during the later phase of the pandemic.

## Data Availability

All data is previousely published data that is cited and referenced in the paper

## Declarations

### Availability of data and material

We analyzed only secondary data available in the public domain.

### Code availability

The code will be made available upon publication in an open GitHub.

### Authors’ contributions

Conceptualization: YS. Analysis: OM. Writing: YS, OM.

### Ethics approval

We analyzed only secondary data available in the public domain, with no need for approval by the ethics committee in research.

